# Inexpensive, versatile and open-source methods for SARS-CoV-2 detection

**DOI:** 10.1101/2020.09.16.20193466

**Authors:** Thomas G.W. Graham, Claire Dugast-Darzacq, Gina M. Dailey, Xammy H. Nguyenla, Erik Van Dis, Meagan N. Esbin, Abrar Abidi, Sarah A. Stanley, Xavier Darzacq, Robert Tjian

**Affiliations:** Department of Molecular and Cell Biology, University of California Berkeley, Berkeley, California 94720, United States; School of Public Health, Division of Infectious Diseases and Vaccinology, University of California Berkeley, Berkeley, California 94720, United States; The Howard Hughes Medical Institute, University of California Berkeley, Berkeley, California 94720, United States

**Author notes:** Equal contribution (Thomas G.W. Graham, Claire Dugast-Darzacq, Gina M. Dailey).

## Abstract

Re-opening of communities in the midst of the ongoing COVID-19 pandemic has ignited a second wave of infections in many places around the world. Mitigating the risk of reopening will require widespread SARS-CoV-2 testing, which would be greatly facilitated by simple, rapid, and inexpensive testing methods. To this end, we evaluated several protocols for RNA extraction and RT-qPCR that are simpler and less expensive than prevailing methods. First, we show that isopropanol precipitation provides an effective means of RNA extraction from nasopharyngeal (NP) swab samples. Second, we evaluate direct addition of NP swab samples to RT-qPCR reactions without an RNA extraction step. We describe a simple, inexpensive swab collection solution suitable for direct addition, which we validate using contrived swab samples. Third, we describe an open-source master mix for RT-qPCR and show that it permits detection of viral RNA in NP swab samples. Lastly, we show that an end-point fluorescence measurement provides an accurate diagnostic readout without requiring a qPCR thermocycler. Adoption of these simple, inexpensive methods has the potential to significantly reduce the time and expense of COVID-19 testing.

## Introduction

The current global pandemic of SARS-CoV-2 has now infected an estimated 29 million people worldwide and claimed over 919,000 lives (Worldometer, https://www.worldometers.info/coronavirus/, accessed 9-11-2020). However, the true number of cases is likely to be even higher, and a full understanding of the scope of the pandemic has been hindered by a persistent lack of widespread testing. The U.S. Centers for Disease Control and Prevention “gold standard” test for COVID-19 detects SARS-CoV-2 viral RNA purified from patient nasopharyngeal swabs. Researchers and clinicians aiming to implement RT-PCR testing for COVID-19 have faced a shortage of the necessary reagents to perform tests in addition to the long processing times required for each test [1]. It has been argued that assays that are less sensitive yet more widely available may be more useful than exquisitely sensitive tests with limited availability [2]. The use of inexpensive, readily procurable reagents and the optimization of rate-limiting steps such as RNA extraction would help to increase the availability of tests and reduce their turnaround time.

Many current RT-PCR protocols for COVID-19 detection, including the CDC-approved test, employ an RNA extraction step to isolate and concentrate viral RNA from patient nasopharyngeal swabs prior to amplification. Typically, this involves the use of a column-based extraction kit such as the Qiagen QIAmp Viral RNA kit or a magnetic bead-based method such as the Roche MagNA Pure kit [3]. Reliance on these commercial kits created supply shortages that hindered testing [4]. Traditional laboratory techniques for RNA purification may offer less expensive alternatives to commercial kits. Trizol extraction followed by isopropanol precipitation provides a high yield of purified RNA [5], however, it requires extensive labor, is difficult to scale to high-throughput, and involves hazardous materials. Simpler isopropanol precipitation methods, in which patient swab samples are first mixed with commercial or homemade lysis solutions, have been reported to give Ct values comparable to those obtained using commercial RNA purification kits [6-8].

To obviate the need for RNA purification altogether, several groups have developed protocols for direct addition of swab samples to RT-qPCRs (reviewed in [9]). While this affords a substantial reduction in the time and expense of testing, the absence of a purification step means that RNA is not concentrated, limiting the sensitivity of detection. Moreover, commonly used swab collection solutions may inhibit RT-PCR. Indeed, while several groups have demonstrated RNA amplification by direct addition of swab samples in the widely used viral transport medium (VTM), inhibition of RT-PCR by VTM typically leads to a significant delay in amplification [10-15]. A comparison of commercial master mixes found that the commonly used TaqPath master mix is particularly susceptible to inhibition by VTM [16].

Consequently, researchers have sought other swab collection solutions compatible with direct addition. Commonly used swab collection solutions, including universal transport medium (UTM), M6, or Hank’s medium, have all been shown to work to some extent for direct addition [17-20]. Some RT-PCR-compatible commercial lysis solutions have also been used to detect SARS-CoV-2 by direct addition [21-23], however the high cost of these products may preclude widespread use. An ideal swab collection solution would be widely available or cheaply made in any laboratory, allow for sensitive, direct detection of patient swabs, and not require specialized storage conditions. Proposed swab collection solutions include saline [18], PBS [14], TE [24], or simply distilled water [15,18]. Also, addition of proteinase K to UTM or saline was reported to improve detection of viral RNA by direct addition [19,25].

Finally, most SARS-CoV-2 testing protocols in clinical use or in pre-clinical development rely on commercial one-step RT-qPCR master mixes [5,13,18,19,26-32]. However, the high cost of commercial master mixes could be prohibitive for widespread testing in resource-limited settings. Master mixes assembled using homemade enzymes may help to address this need [33-35].

Here, we show that a simple isopropanol precipitation protocol provides an effective means of extracting RNA from nasopharyngeal (NP) swab samples that is suitable for subsequent RT-qPCR detection. As an alternative approach, we evaluated direct addition of swab samples to RT-qPCRs. Consistent with previous reports, we find that while direct addition of small amounts of swab sample in UTM permits SARS-CoV-2 detection, inhibition of the reaction by UTM limits the amount of sample that can be added, and hence the detection sensitivity. We evaluate alternative swab collection media and find that a simple solution of proteinase K in water permits sensitive detection of RNA from in vitro-cultured SARS-CoV-2 in contrived swab samples containing human nasal mucus. Finally, we describe “BEARmix”, a one-step RT-qPCR master mix that can be assembled using homemade Taq polymerase and M-MLV reverse transcriptase. This master mix is substantially (~80x) less expensive than commercial master mixes and permits the detection of as few as tens of RNAs per reaction. Taken together, homemade methods such as these have the potential to circumvent reliance on commercial kits and reagents, dramatically lower the cost per test, and facilitate widespread testing.

## Results

### Isopropanol precipitation of RNA for SARS-CoV-2 detection

As an alternative to commercial RNA purification kits, we evaluated a simple isopropanol precipitation procedure using inexpensive components (see Materials and methods). When tested using a mixture of human cell RNA and in vitro-transcribed SARS-CoV-2 N gene RNA, isopropanol precipitation gave RNA recovery comparable to the QIAmp Viral kit and significantly better than the Qiagen RNeasy Mini Kit (Fig 1A).We next evaluated isopropanol precipitation using the same NP swab samples described in a previous publication [26,36]. Swab samples in universal transport medium (UTM) were inactivated either by addition of 1 volume of 2x DNA/RNA Shield (samples Neg1-Neg2 and Pos1-Pos2), or by treatment with 0.4 mg/ml proteinase K for 30 min at 37°C, followed by heat-inactivation at 95°C for 5 min and 75°C for 30 min (Neg3-Neg12 and Pos3-Pos12). RNA was then purified using either the Qiagen RNeasy Mini kit or isopropanol precipitation (see Materials and methods). Due to supply shortages, the QIAmp Viral RNA extraction kit was not available when we performed these experiments. Purified RNA was then assayed using Thermo Fisher TaqPath master mix with the CDC-approved N1, N2 and RNase P probe sets. 11 out of 12 known positive samples showed amplification using isopropanol-precipitated RNA, while only 9 out of 12 positive samples showed amplification using Qiagen RNeasy-purified RNA (Fig 1C). The sample that failed to show amplification by both methods, Pos5, had also shown very high Ct values using magnetic-bead based RNA purification (see Fig 5A in [36]). The relatively poor performance of the Qiagen RNeasy kit is consistent with a previous report showing that this kit gives higher Cq values than the CDC-recommended QIAamp Viral RNA extraction kit [17]. None of the negative samples showed amplification of viral RNA using either extraction method, while all positive and negative samples (with the exception of sample Pos5 for the RNeasy kit) showed amplification using the human RNase P positive control probe set (Fig 1B-C).

**Fig 1.**
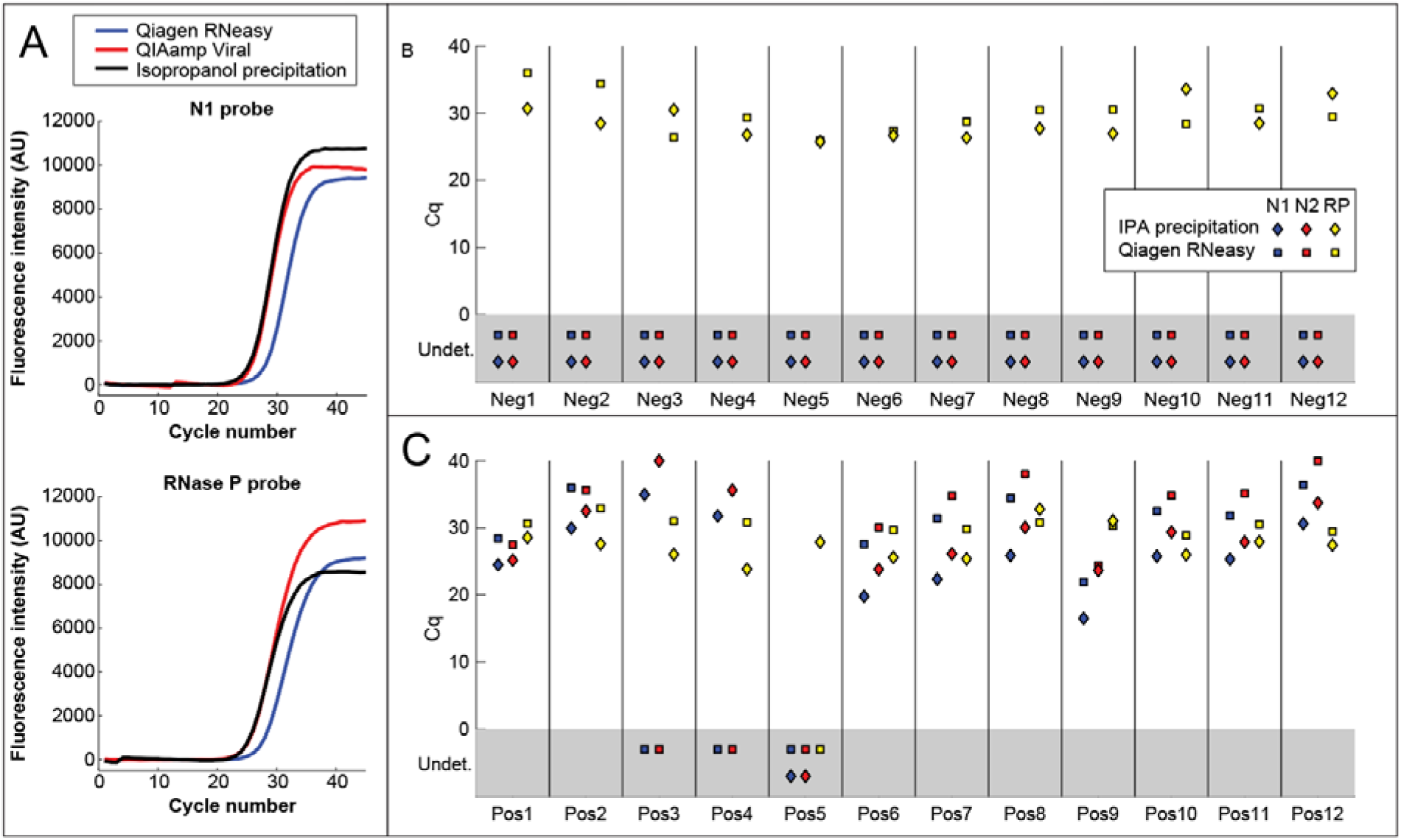
RNA extraction by isopropanol precipitation. (A) TaqPath amplification curves of a mixture of human, cell RNA and in vitro-transcribed SARS-CoV-2 N gene RNA purified using the Qiagen RNeasy Mini kit, QIAamp Viral RNA Mini kit, or isopropanol precipitation. Isopropanol precipitation gives RNA recovery better than the RNeasy kit and comparable to the QIAamp Viral kit. (B-C) Cq values of RNA from positive (Pos) and negative (Neg) NP swab samples in UTM, purified using either the Qiagen RNeasy Mini Kit (squares) or isopropanol precipitation (diamonds). Reactions were performed using TaqPath master mix with the CDC SARS-CoV-2 N1 and N2 probes (blue and red points) and the human RNAse P (RP) control probe (yellow points). For points in the gray rectangle, no amplification was observed and Cq values were “undetermined” (Undet.).

### Direct addition of swab samples to RT-PCR reactions

Potentially more useful than simplifying RNA purification would be foregoing RNA purification entirely (see Introduction). We therefore tested whether we could detect SARS-CoV-2 RNA by adding 1 µl of each swab sample to 20 µl TaqPath reactions containing the N1, N2, and RNase P (RP) probes (Fig 2A). In the interest of safety, we first heat-inactivated aliquots of samples Pos1-Pos2 and Neg1-Neg2 under BSL3 conditions using three different protocols: 1) 75°C for 30 min, 2) 95°C for 5 min, followed by 75°C for 30 min, 3) 30 min at 37°C in the presence of 0.4 mg/ml proteinase K, followed by 95°C for 5 min and 75°C for 30 min. Each protocol includes at least 30 min at 75°C, as this was found to be sufficient to inactivate the virus (S3 Fig and data not shown). Amplification with primers N1 and N2 was observed for both positive samples (Fig 2A). Neither negative sample showed amplification with primer sets N1 and N2, and RNase P amplification was observed for all samples. Because Protocol 3 gave slightly lower Cq values for viral RNA than Protocols 1 and 2 (Fig 2A), we proceeded to heat-inactivate aliquots of the remaining positive and negative NP swab samples using Protocol 3. We then tested each sample by direct addition of 1 µl to 20 µl TaqPath reactions with N1, N2, and RP probes. Amplification was observed using both N1 and N2 in 10 out of 12 positive samples, and 0 out of 12 negative samples (Fig 2A-B). Sample Pos3, which had the highest Cq values for N1 and N2 using the isopropanol precipitation method, showed very late amplification with N2 but not with N1. All samples showed amplification using the RNase P control probe. Thus, results from direct addition of 1 µl of swab sample were concordant in most cases with results from the standard RNA purification-based assay.

**Fig 2.**
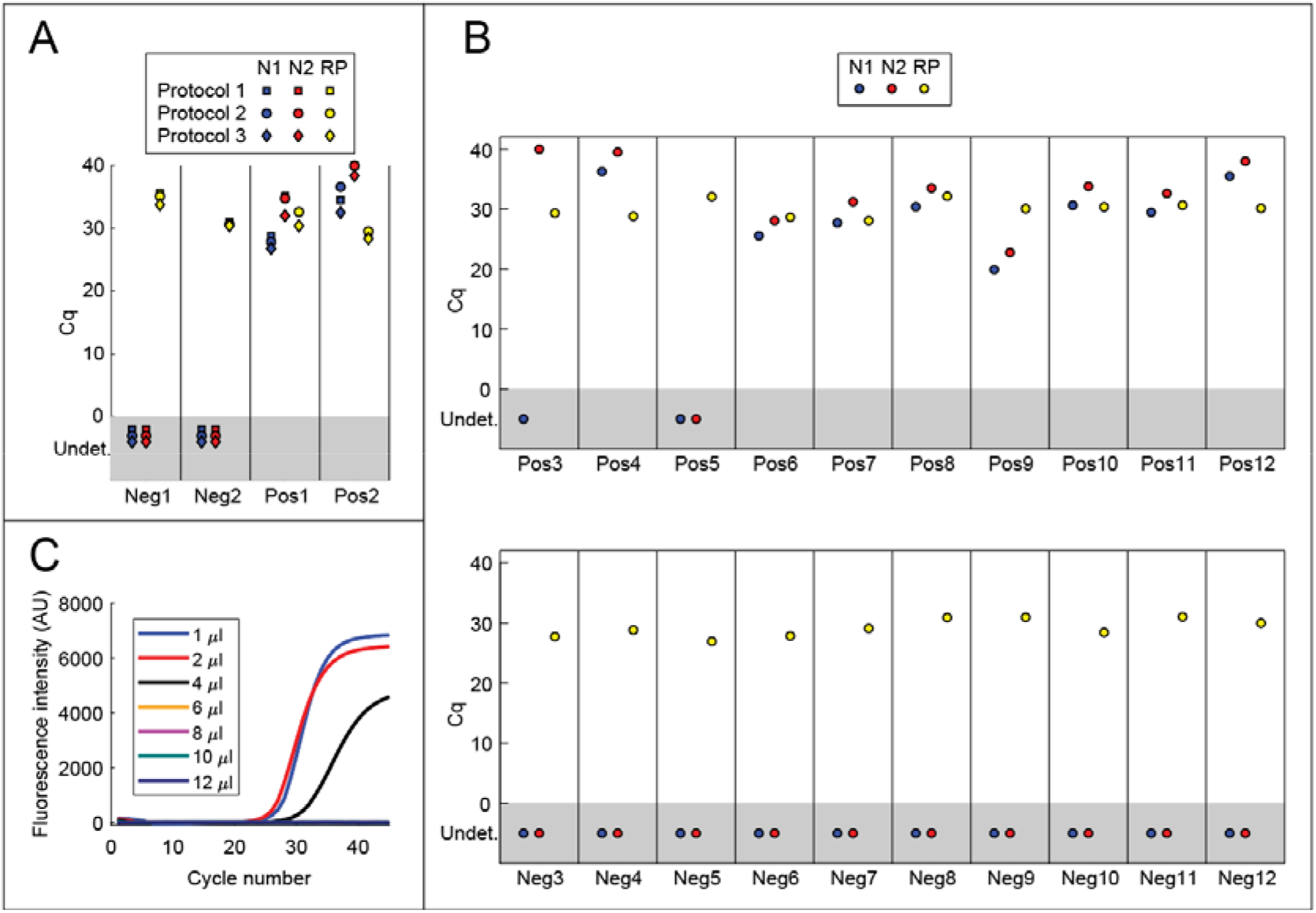
Direct addition of swab samples to RT-PCRs. (A) Direct addition of 1 µl of swab sample to 20 µl TaqPath reactions containing N1 (blue), N2 (red), or RNase P (RP; yellow) primer/probe mixtures. Samples were heat-inactivated using one of three protocols: 1) 75°C for 30 min, 2) 95°C for 5 min, followed by 75°C for 30 min, 3) 37°C for 30 min in the presence of 0.4 mg/ml proteinase K, followed by 95°C for 5 min and 75°C for 30 min. (B) Detection of SARS-CoV-2 in positive (Pos) and negative (Neg) NP swab samples by direct addition. Proteinase K was added to each sample to a final concentration of 0.4 mg/ml, and samples were incubated at 37°C for 30 min, 95°C for 5 min and 75°C for 30 min. 1 µl of swab sample was added to 20 µl TaqPath reactions containing N1 (blue), N2 (red), and RNase P (RP; yellow) primer/probe mixtures. (C) Direct addition of different quantities of heat-inactivated swab samples in UTM to TaqPath master mix. The indicated amounts of positive swab sample Pos1 were added to 20 µl TaqPath reactions containing probe N1. See also Fig. S1.

To test whether direct addition of larger quantities of swab sample would yield lower Cq values, we compared addition of 1 µl, 2 µl, 4 µl, 6 µl, 8 µl, 10 µl, and 12 µl of sample to a 20 µl TaqPath reaction. Despite the addition of twice as much RNA, Cq values were similar with 2 µl of sample as with 1 µl (Fig 2C and S1 Fig). Furthermore, amplification was inhibited by 4 µl or greater of swab sample. Taken together, our results confirm that viral RNA may be detected by direct addition of swab samples in UTM to TaqPath master mix, as long as the amount of swab sample added does not exceed ~5-10% of the total reaction volume.

### Development of an alternative swab collection solution compatible with RT-PCR

We reasoned that collecting swab samples in a buffer that does not inhibit RT-qPCR would permit the addition of a greater volume of swab sample per reaction, potentially increasing the sensitivity of the assay. To this end, we first tested whether various additives inhibit TaqPath master mix when 5 µl of each are added to a 10 µl reaction (S2 Fig). Minimal inhibition of TaqPath (ΔCq < 0.5) was seen by 1x TE, 10 mM Tris (pH 7.5, 8, 8.3, or 8.5), ≤ 2 mM EDTA, ≤ 0.5% Triton X-100, ≤ 2% NP-40, ≤ 2% Tween-20, ≤ 2% ICA-630, ≤ 0.02% Sarkosyl, or ≤ 10 mM DTT. Slight inhibition was observed for ≥ 1% Triton X-100 and 0.05% Sarkosyl, while complete inhibition was observed for 1x PBS and ≥ 0.2% Sarkosyl.

Based on these results, we prepared two candidate solutions containing non-inhibitory components— Tris-HCl, pH 8, dilute EDTA, Tween-20, and DTT—and prepared 20 μl TaqPath reactions containing 10 μl of in vitro-transcribed N gene RNA diluted in either these solutions or water. Both solutions gave comparable Cq values to water at each RNA concentration, indicating that both are compatible with direct addition to TaqPath master mix (Fig 3A).

**Fig 3.**
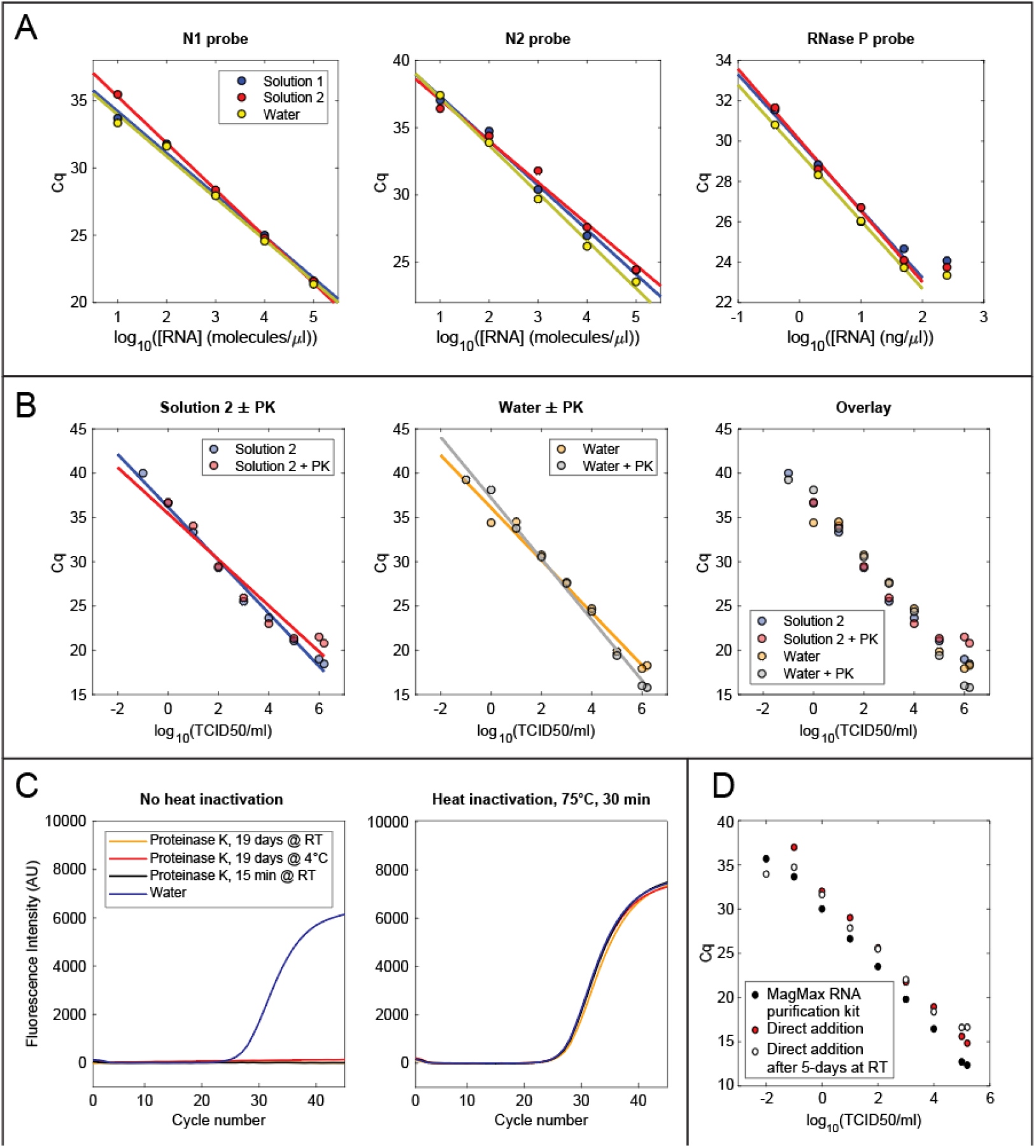
Swab collection solutions optimized for direct addition. (A) RT-qPCR of N gene RNA or human cell RNA in swab collection solutions. RNA was diluted to the indicated concentration in Solution 1 (10 mM Tris, pH 8, 1 mM EDTA, 0.2% Tween 20, 0.2 mM DTT), Solution 2 (10 mM Tris, pH 8, 1 mM EDTA, 0.5% Tween 20, 0.5 mM DTT), or water, and 10 µl of each dilution were analyzed in 20 µl TaqPath reactions containing the indicated probes. (B) Direct addition to RT-qPCR of cultured SARS-CoV-2 heat-inactivated with or without proteinase K treatment in either water or Solution 2 (10 mM Tris, pH 8, 1 mM EDTA, 0.5% Tween 20, 0.5 mM DTT). 13.5 µl of each sample was added to a 20 µl TaqPath reaction. (C) Amplification curves of 20 µl TaqPath reactions containing 2 x 10^4^ in vitro-transcribed N gene RNAs, the N2 primer/probe mixture, and 10 µl of proteinase K stored under different conditions. PK solution was either added directly to the reaction (left panel) or heat inactivated first for 30 min at 75°C (right panel). Non-heat-inactivated PK completely inhibited amplification, even after 19 days of storage at room temperature (left panel). However, amplification was not inhibited when PK was heat-inactivated prior to addition (right panel). (D) Comparison of viral detection by direct addition or RNA extraction with the MagMax Viral RNA isolation kit (Thermo Fisher). Cultured SARS-CoV-2 was diluted to the indicated number of infectious units into 0.4 mg/ml proteinase K in water. RNA was analyzed using TaqPath master mix and the N1 primer/probe mixture, either by direct addition of 13.5 µl of heat-inactivated sample to a 20 µl reaction or by addition of 5 µl of purified RNA to a 20 µl reaction.

To evaluate detection of actual virus by direct addition to an RT-qPCR, we prepared serial dilutions of in vitro-cultured SARS-CoV-2 in 1x PBS and mixed 1 volume of each dilution with 9 volumes of either water or buffer 2. Because proteinase K treatment gave lower Cq values for NP swab samples, we also prepared the same mixtures with 500 μg/ml proteinase K. All samples were incubated at 37°C for 30 min and heat-inactivated at 75°C for 30 min. Complete heat-inactivation of virus in each solution was confirmed using cytopathic effect assays (CPE) in Vero E6 cells (S3 Fig, see Materials and methods). For a given viral dilution, similar Cq values were obtained in all four solutions (Fig 3B). In contrast to NP swab samples, treatment with 500 μg/ml PK did not reduce the Cq values for direct addition of cultured virus (see Discussion).

We next compared our direct addition protocol to the protocol used by the CLIA-approved SARS-CoV-2 testing center at the UC Berkeley Innovative Genomics Institute, which relies on the Thermo Fisher MagMax RNA purification kit [26]. Because RNA is concentrated 18-fold by this purification procedure, it is expected that amplification would be delayed by approximately 2.7 cycles in a direct-addition reaction with 13.5 µl of unconcentrated sample compared to a reaction with 5 µl of purified RNA. Notably, we observed an average delay of 2.5 cycles for the N1 primer (range 2.0 to 3.3), implying that amplification of RNA from crude, heat-inactivated virus is approximately as efficient as amplification of an equivalent amount of purified RNA (Fig 3D).

Because we and others have found that proteinase K improves RNA extraction from swab samples, we evaluated the shelf-life of PK dissolved in water or Solution 2. PK was diluted to 200 jg/ml in either water or Solution 2 and stored for up to 19 days at room temperature or 4°C. A BSA proteolysis assay was used to measure PK activity either immediately after dilution or after 1, 5, 12, or 19 days of storage (see Materials and methods). PK remained active after 19 days of storage in water at either room temperature or 4°C (S4 Fig). By contrast, PK stored in Solution 2 showed reduced activity after 19 days of storage at room temperature (S4 Fig). These experiments demonstrate that PK may be stored in Solution 2 at 4°C or in water at either 4°C or room temperature for over 2 weeks with no measurable loss of activity.

### An open-source, one-step RT-qPCR master mix for SARS-CoV-2 detection

In addition to RNA purification kits, commercial RT-qPCR master mixes are an expensive testing component. We therefore sought to develop a one-step RT-qPCR master mix consisting of homemade, off-patent enzymes and inexpensive buffer components. After evaluating various enzymes and buffers, we achieved the most consistent results by using a combination of M-MLV reverse transcriptase (specifically, the RNase H-deficient D524N mutant [37]) and Taq polymerase in a buffer containing a high concentration of trehalose. We call this mixture BEARmix (basic economical amplification reaction mix). A 4x mixture of BEARmix buffer and enzymes lost activity when it was stored at −20°C and subjected to multiple freeze-thaw cycles (data not shown). Thus, we recommend that the enzymes and buffer + dNTP mixture be stored separately and combined just before reaction setup.,

Using dilutions of in vitro-transcribed SARS-CoV-2 N gene RNA and the N2 primer/probe set, we observed the expected log-linear relationship between Cq value and amount of input RNA (Fig 4A, upper panel). Amplification was consistently observed with as few as 50 RNAs per reaction, however a lack of amplification was sometimes observed fo,r lower quantities of RNA (Fig 4A, lower panel and S5 Fig). Raw fluorescence traces without background subtraction exhibited a slow linear increase in baseline fluorescence, even in the absence of template (S6 Fig), which may arise from slow degradation of free probe oligonucleotides by the enzyme mixture. Cq values were thus determined based on the second derivative of fluorescence intensity, which is unaffected by the addition of linear baseline drift ([38]; see Materials and methods and S6 Fig).

**Fig 4.**
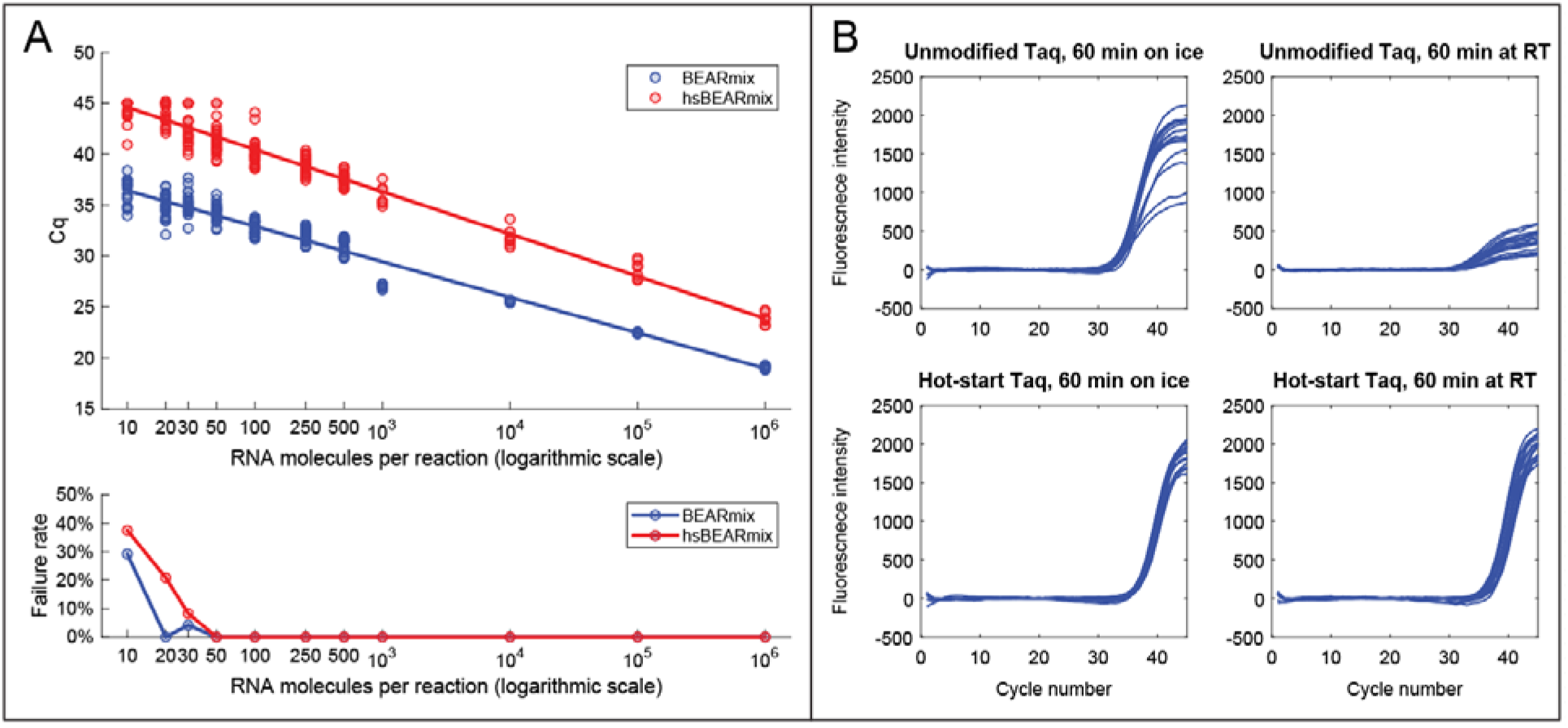
“BEARmix”: an open-source, one-step RT-qPCR master mix. (A) Top panel: Standard curves of BEARmix with unmodified Taq polymerase (blue) and formaldehyde-crosslinked hot-start Taq polymerase (red). Linear fit for non-hot-start BEARmix: Cq = 39.9 - 3.489x (amplification factor 1.93), where x is the logarithm with base 10 of the number of RNAs perreaction. Linear fit for hot-start BEARmix: Cq = 48.8 - 4.15x (amplification factor = 1.74). Bottom panel: Fraction of reactions that did not show amplification (failure rate) for each RNA concentration. N = 24 reactions for 10 to 500 copies of RNA, N = 8 reactions for 1000 to 106 copies. (B) Homemade hot-start Taq polymerase permits reaction setup at room temperature. BEARmix reactions were set up using unmodified and hot-start (crosslinked) Taq polymerase with 20 molecules of N gene RNA per reaction. Reactions were performed in a qPCR thermocycler after incubation for 60 min either on ice or at room temperature. In contrast to regular Taq polymerase, amplification by hot-start Taq polymerase is not inhibited by incubating reactions for 60 min at room temperature prior to running the RT-qPCR cycle.

A drawback of wild-type Taq polymerase is that it can extend mispaired primers at room temperature, producing “primer dimer” products that compete for amplification with the target amplicon [39-41]. To overcome this problem, companies have generated “hot-start” versions of Taq polymerase, typically by including a proprietary monoclonal antibody or aptamer in the reaction, which inhibits the polymerase at low temperatures but is denatured at high temperature [39-41]. Because these approaches are expensive or patent-protected, we evaluated an off-patent method for producing hot-start Taq polymerase using formaldehyde fixation [42-44]. Treatment with formaldehyde produces crosslinks within the enzyme that inhibit its activity, while incubation at 95°C during the PCR cycle reverses the crosslinks to restore enzymatic activity. We prepared formaldehyde-crosslinked hot-start Taq polymerase and compared it with non-crosslinked Taq polymerase in reactions with N gene RNA and the N1 primer/probe set. Reactions were incubated either on ice or at room temperature for various lengths of time after primer addition. Reactions containing unmodified Taq polymerase showed substantially reduced amplification after a 10-minute incubation at room temperature, and amplification was drastically reduced after 1 hour at room temperature (Fig 4B, top row). In contrast, reactions containing hot-start Taq polymerase showed amplification even after a 1-hour incubation at room temperature (Fig 4B, bottom row). However, amplification of a given quantity of RNA with hot-start Taq polymerase occurred at a later cycle than with regular Taq polymerase (Fig 4A-B). This was the case across a wide range of input RNA concentrations (Fig 4A), which may reflect incomplete reactivation of the enzyme by heating at 95°C. Consistent with this interpretation, the amplification efficiency, as judged by the slope of the Cq vs. RNA concentration curve, was lower for hot-start Taq than for regular Taq (amplification factor of 1.74 for hot-start Taq versus 1.93 for unmodified Taq). Increasing the time of the 95°C uncrosslinking step to 15 or 20 minutes led to earlier amplification, however amplification with crosslinked Taq was still delayed relative to uncrosslinked Taq (S7 Fig). Thus, formaldehyde crosslinking of Taq permits reaction setup at room temperature, albeit with reduced amplification efficiency.

We used BEARmix to perform RT-qPCR on the remaining isopropanol-precipitated RNA from the NP swab samples that we had analyzed previously with TaqPath master mix (Fig 5A). Amplification with both N1 and N2 probes was observed in 6 out of 9 positive samples tested, and amplification was observed with N2 but not N1 for sample Pos12. Amplification was not observed with either N1 or N2 for samples Pos3 and Pos4, which previously had the highest Cq values with TaqPath master mix. None of the negative samples showed amplification with N1 or N2, while all positive and negative samples showed amplification with the RNase P control probe. Direct addition of 0.5 µl of swab samples to 10 µl BEARmix reactions gave amplification in at least one replicate for 10/12 positive samples and 0/12 negative samples (Fig 5B). However, amplification failed for at least one replicate in three positive samples, while samples Pos3 and Pos4 failed to show amplification in any replicates. Taken together, these results show that RT-qPCR with BEARmix can detect SARS-CoV-2 in clinical samples, either using purified RNA or by direct addition of swab samples, albeit with somewhat less sensitivity than commercial TaqPath master mix. It is conceivable that sample degradation contributed to the observed reduction in sensitivity, as RNA samples were frozen after being assayed with TaqPath, stored at −80°C for 1 week, and thawed for testing with BEARmix.

**Fig 5.**
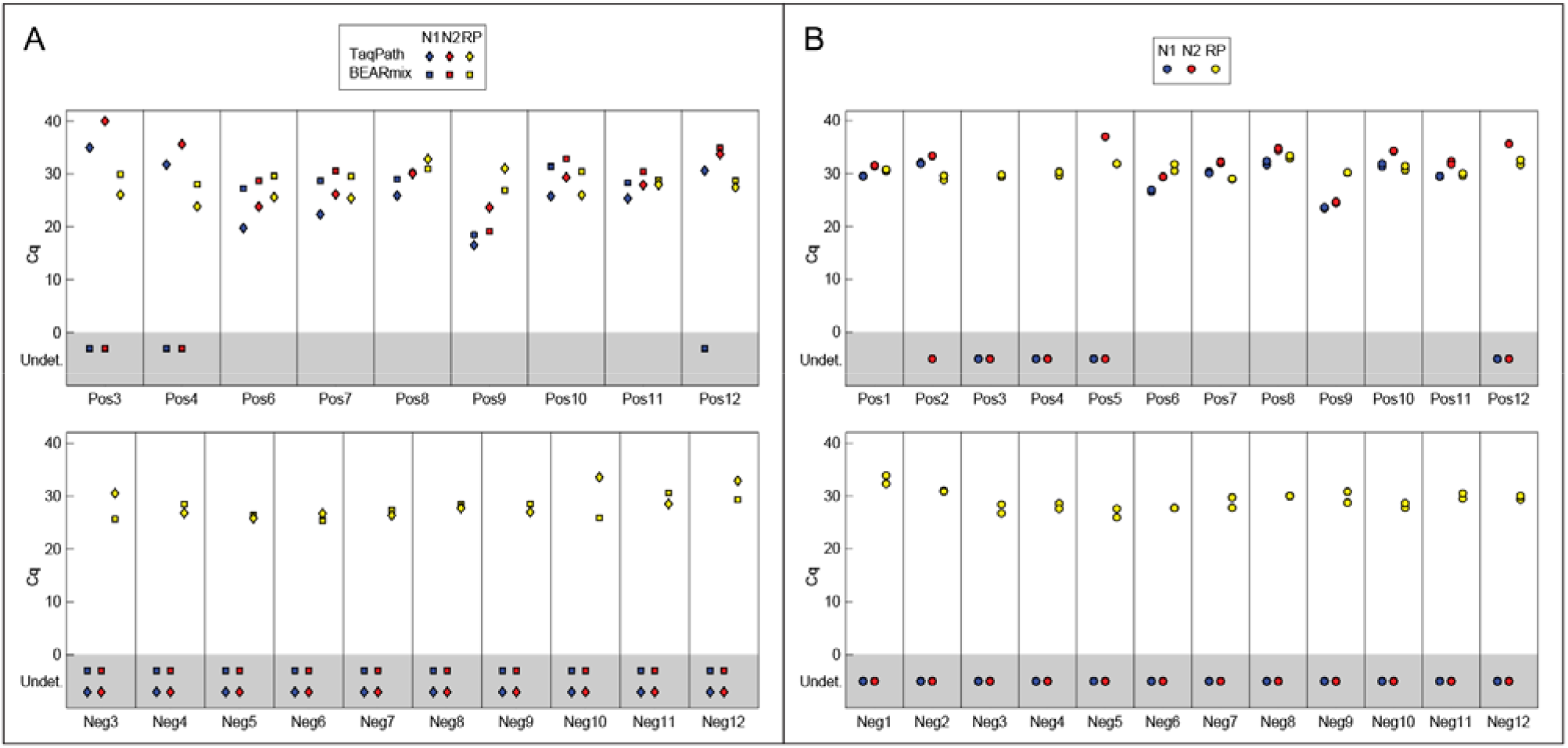
Analysis of clinical samples using BEARmix. (A) Cq values from RT-qPCR of isopropanol-precipitated NP swab samples using BEARmix. For comparison, TaqPath Cq values for the same samples are re-plotted from Fig. 1B-C. (B) Direct addition of 0.5 µl of clinical swab samples in UTM to 10 µl BEARmix reactions. Pairs of points represent qPCR duplicates.

### Analysis of contrived swab samples by direct addition to an open-source master mix

To evaluate a complete protocol in which swab samples are collected into PK solution and then added directly to BEARmix RT-PCRs, we prepared contrived swab samples in which live virus was mixed with pathogen-free human nasal fluid prior to dilution into either DNA/RNA Shield, VCM containing 0.4 mg/ml proteinase K, or a solution of 0.4 mg/ml proteinase K in water (Fig 6). Samples in water + PK and VCM + PK were incubated for 30 min at 37°C and then heat-inactivated at 75°C for 30 min.

**Fig 6.**
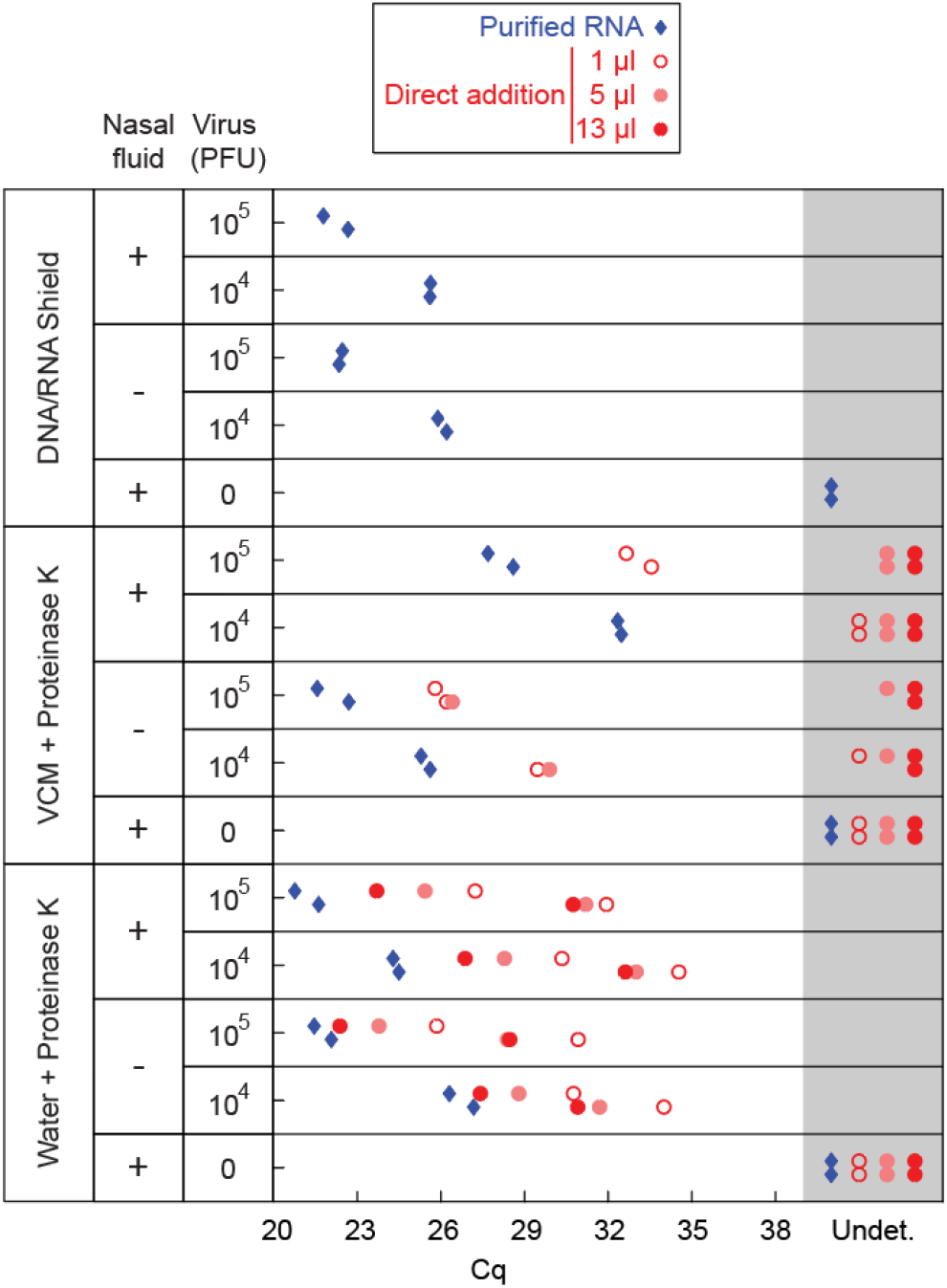
RT-qPCR of contrived swab samples. Indicated quantities of in vitro cultured SARS-CoV-2 were mixed with the swab collection solutions listed in the leftmost column, either alone or in combination with human nasal fluid. Samples were analyzed by RT-qPCR using BEARmix with the N1 primer/probe set either after RNA extraction with the QIAmp Viral RNA purification kit (blue diamonds) or by direct addition (red circles). Two qPCR replicates are shown in separate vertical rows for each condition.

To assess RNA integrity, we extracted viral RNA from all of the samples using the QIAmp Viral RNA extraction kit and performed RT-qPCR using the N1 primer/probe mixture. The presence of nasal fluid did not inhibit RNA amplification for samples in DNA/RNA Shield or water + PK (Fig 6, blue diamonds), indicating that viral RNA is preserved in PK solution in the presence of nasal fluid. However, higher Cq values were observed in the presence of nasal fluid in VCM + PK, suggesting that RNA is not preserved as well in this solution in the presence of nasal fluid.

Next, we evaluated amplification of contrived swab samples added directly to RT-qPCRs. As with samples in UTM (Fig 2C), addition of more than 1 µl of contrived swab sample in VCM + PK did not give lower Cq values, but instead inhibited amplification (Fig 6, middle section, red circles). In contrast, addition of increasing amounts of contrived swab sample in water + PK led to lower Cq values (Fig 6, bottom section, red circles). As expected, Cq values were higher for direct addition of contrived swab samples than for purified, concentrated RNA. Thus, while direct addition of swab samples in PK solution provides somewhat lower sensitivity than addition of purified, concentrated RNA, the option to add a larger volume of samples in PK solution improves detection relative to samples in VCM.

### Endpoint detection with a fluorescence imager

Real-time qPCR thermocyclers are expensive instruments, which some testing centers have had to borrow from academic labs [26]. However, standard thermocyclers are relatively inexpensive and ubiquitous, and even less expensive, miniaturized PCR machines have been developed [45]. Thus, we tested whether it is possible to distinguish the presence or absence of viral RNA without a real-time fluorescence readout by visual inspection of images from a standard fluorescence gel imager. We imaged RT-qPCR plates from the experiments in Fig 2B and 1B-C on a BioRad Chemidoc imager in the fluorescein (TaqMan probe) and rhodamine (loading control) channels (Fig 7A-B). Wells that had shown amplification in real-time traces exhibited visibly greater fluorescence in the fluorescein channel from those that had not (Fig 7A-B). Similarly, imaging in the fluorescein channel clearly distinguished BEARmix reactions containing 100 or 1000 in vitro transcribed N gene RNAs from negative control reactions without RNA (Fig 7C). Given that the goal of testing is a binary determination of whether patients are positive or negative for the virus, an endpoint assay of this sort could potentially provide the desired information without an expensive real-time PCR instrument.

**Fig 7.**
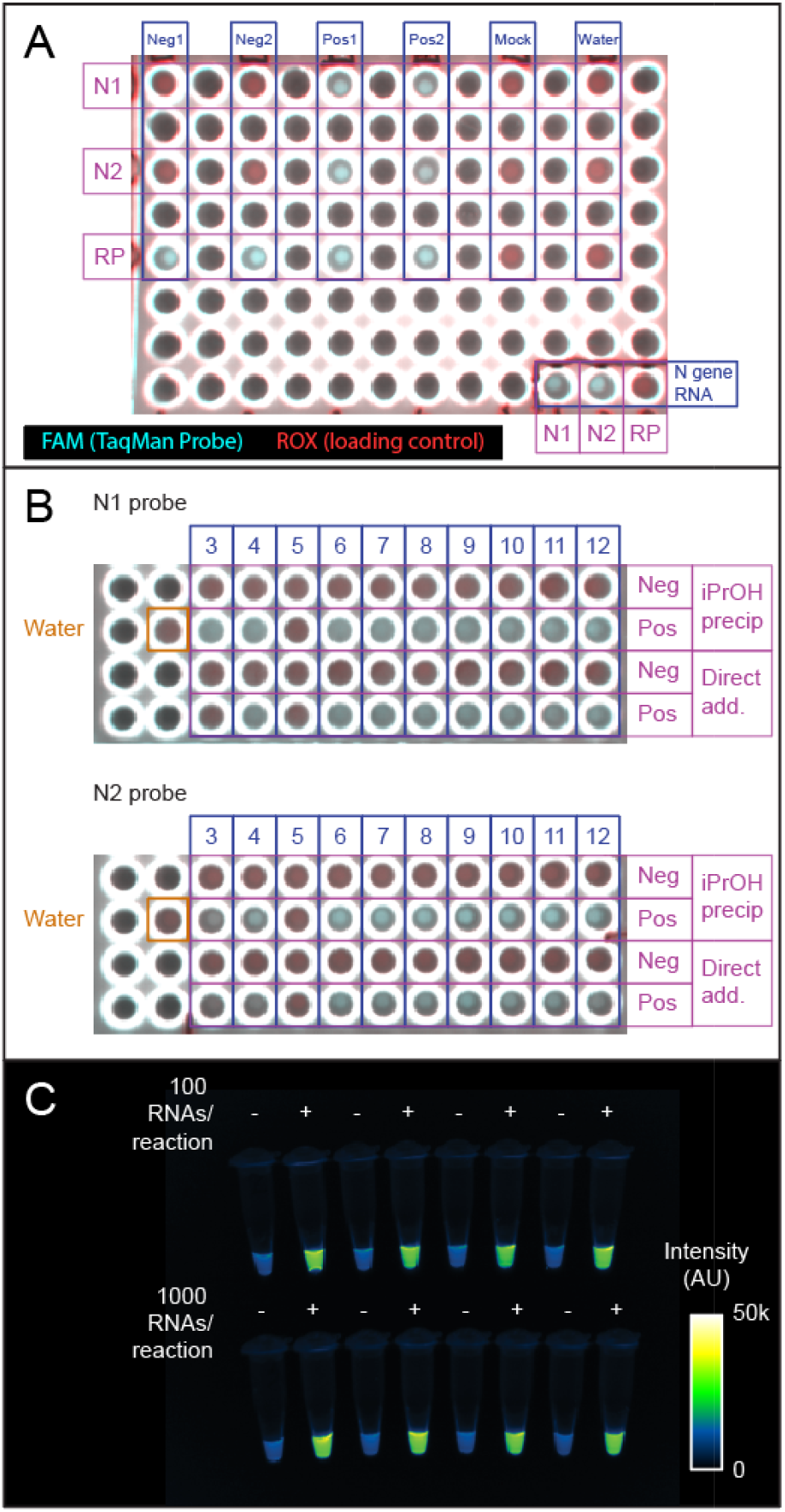
Endpoint fluorescence detection. (A) Endpoint fluorescence image of the qPCR plate used for the first two clinical samples in Fig. 1B-C. Shown is a 2-channel overlay in which the ROX control dye in TaqPath master mix appears in the rhodamine channel (red) and dequenched FAM product from the TaqMan probe appears in the fluorescein (cyan) channel. An N gene RNA positive control is in the lower right-hand corner. Positive and negative samples are clearly distinguishable based on fluorescence in the FAM channel. B) BioRad Chemidoc fluorescence image of the qPCR plate used for the IPA precipitation and direct addition reactions in Fig.1B-C and 2B. Positive and negative samples distinguishable by qPCR are also distinguishable by endpoint fluorescence imaging. Red, rhodamine (0.5 s exposure). Cyan, fluorescein (0.05 s exposure). Scale is set from 0 to 55000 counts for each channel. C) Endpoint detection using the N2 probe set and BEARmix. Reactions were set up in alternating tubes with water (negative control) or in vitro transcribed N gene RNA at 100 or 1000 copies per reaction. An image taken with a 0.1 s exposure time in the fluorescein channel of a ChemiDoc imager (BioRad) is displayed using the Fiji “Green Fire Blue” colormap with lower and upper limits set to 0 and 50000.

## Discussion

Here we have developed simple, academic laboratory-derived methods for RNA extraction, direct sample addition, and RT-PCR detection that provide low-cost alternatives to the use of commercial kits (Fig 8). Adoption of these methods may facilitate widespread testing in a cost-effective way.

**Fig 8.**
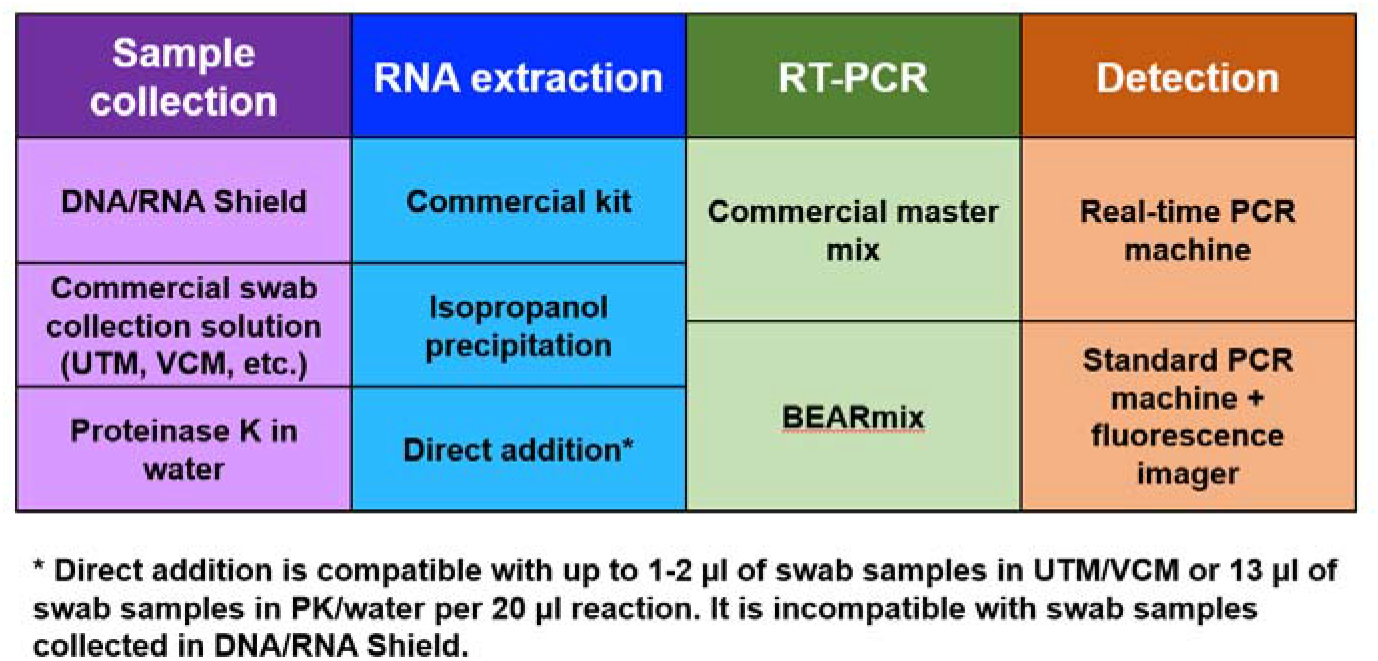
Summary of testing options. Working protocols can be assembled from various combinations of sample collection, RNA extraction, RT-PCR, and detection methods. For each step, we have demonstrated economical alternatives to the prevailing method.

Isopropanol precipitation is an extremely simple and inexpensive method for extracting and concentrating RNA, which provides similar yield to sophisticated commercial kits (Fig 1A), and is somewhat easier and faster in our experience than the use of individual spin columns. Even when applied to complex NP swab samples, it yields RNA suitable for downstream amplification by RT-qPCR (Fig 1B-C). We chose to concentrate RNA between 2-fold and 8-fold, but greater fold concentration could likely be achieved by increasing the amount of input swab sample or decreasing the volume in which the pellet is redissolved. While the throughput of this method may be limited if performed using microcentrifuge tubes, precipitating samples in 96-well plates and removing the supernatant using a multi-well aspirator might allow for a greater number of samples to be processed in parallel.

Direct addition of swab samples to RT-qPCR reactions saves money and time by foregoing an RNA purification step. Consistent with previous studies, we find that it is possible to detect virus by adding a small volume of heat-inactivated swab sample in UTM to an RT-qPCR (Fig 2). Incubation of swab samples with proteinase K prior to heat-inactivation yielded slightly lower Cq values for detection (Fig 2A). Interestingly, this beneficial effect of PK treatment was not observed for cultured virus (compare Fig 2A and 3B), perhaps reflecting degradation by PK of RNases or some other inhibitory protein component that is present in human fluids but not in cell culture supernatant. Unfortunately, inhibition of RT-qPCR by the commonly used swab collection solutions UTM and VCM limits the amount of sample that can be added to the reaction, and hence the sensitivity of detection (Fig 2C and 6). Our results suggest that direct addition would be facilitated by collecting swabs in either a low-salt buffer or water containing proteinase K. Strikingly, direct addition of heat-inactivated virus in low-salt buffer or water gave Cq values close to those expected based on the total RNA copy number, indicating that RT-qPCR amplification is approximately as efficient with heat-inactivated virus as with purified viral RNA (Fig 3D). The direct addition method was likewise effective for contrived swab samples containing cultured virus and human nasal fluid in water + PK, and adding a larger volume of sample generally gave lower Cq values (Fig 6). This method is also safe to use, as the virus can be heat-inactivated without opening the tube (Fig S3). Even with improved collection buffers, there is of course a limit to the volume of sample that can be added per reaction, and direct addition is thus less sensitive than purification methods that concentrate RNA. Testing centers must therefore judge whether reduced sensitivity is worth the time and cost savings of the direct addition method.

Commercial master mixes for one-step RT-qPCR cost up to hundreds of U.S. dollars per milliliter. We describe “BEARmix”, a simple laboratory-derived master mix capable of detecting tens of RNA molecules per reaction. BEARmix is made using M-MLV reverse transcriptase and Taq polymerase, which are easy to purify with high yield in any laboratory equipped for protein biochemistry. A hot-start version of BEARmix can be made by formaldehyde crosslinking Taq polymerase, however this comes with the drawback of less efficient amplification (Fig 4A-B and S7 Fig). BEARmix successfully detected SARS-CoV-2 RNA in a majority of NP swab samples, albeit with generally higher Cq values than commercial TaqPath master mix (Fig 5). It is unclear whether the poorer performance of BEARmix than commercial TaqPath master mix in our experiments reflects an inherent difference between the two mixes, or whether it was due to the greater age of the samples when we tested them with BEARmix. Validating BEARmix for clinical diagnostics would require a side-by-side comparison of BEARmix and a commercial master mix using a larger number of recently collected patient samples. Additionally, it would be interesting to evaluate BEARmix in combination with recently developed direct-addition protocols for saliva testing [46,47]. We can also imagine various ways of improving our master mix, for instance, by including dUTP and UDG to prevent amplicon contamination, optimizing the conditions for hot-start Taq preparation and reactivation, or testing other public-domain DNA polymerase and reverse transcriptase variants [48].

Expanding the scope of SARS-CoV-2 testing will be crucial to contain the pandemic until treatments and vaccines are available. High frequency and low turnaround time of testing may be even more critical than high sensitivity. Our results demonstrate that relatively simple and inexpensive methods can be used to detect SARS-CoV-2 with reasonable sensitivity. Continued refinement of these methods and their adoption by “pop-up” testing centers [26,46,47] could facilitate expanded testing.

## Materials and methods

### In vitro transcription of N gene RNA

The SARS-CoV-2 N gene sequence was amplified from the Integrated DNA Technologies (IDT) N gene control plasmid (Cat. # 10006625) using primers T7_nCoV_N_F (5’

TAATACGACTCACTATAGGGatgtctgataatggaccccaaaatc 3’) and M13R (5’ caggaaacagctatgaccatg 3’). This was gel-purified using the Zymoclean Gel DNA Recovery Kit (Zymo Research), and ~70 ng of gel-purified PCR product was in vitro transcribed using the HiScribe T7 Quick Kit (NEB) in a 20 µl reaction.

Following overnight incubation at 37°C, RNA was purified using the Qiagen RNeasy kit. The concentration of the purified RNA was determined on a NanoDrop spectrophotometer and converted into molar concentration using a calculated molecular weight of 4.2 x 10^5^ g/mol. RNA was diluted to a working stock concentration of 10^6^ molecules per µl, aliquotted, and stored at −80°C.

### Obtaining samples

De-identified nasopharyngeal (NP) swab samples positive and negative for SARS-CoV-2 were obtained from Kaiser Permanente Healthcare, as described in a previous publication [26].

### Inactivation with DNA/RNA Shield

For chemical inactivation, NP swab samples and samples of cultured virus were combined with an equal volume of 2x DNA/RNA Shield (Zymo Research) under BSL3 conditions, mixed thoroughly, and incubated for 20 min at room temperature. Samples were then transferred to new vials prior to being transported out of the BSL3 facility.

### Heat-inactivation and cytopathic effect (CPE) assays

NP swab samples in UTM were heat-inactivated under BSL3 conditions using one of three protocols: 1) 75°C for 30 min, 2) 95°C for 5 min, followed by 75°C for 30 min, 3) 37°C for 30 min in the presence of 0.4 mg/ml proteinase K, followed by 95°C for 5 min and 75°C for 30 min. Samples of cultured virus were inactivated by incubating for 37°C for 30 min (either with or without 0.5 mg/ml proteinase K) followed by 75°C for 30 min. Inactivated samples were subsequently transferred to new vials prior to being transported out of the BSL3 facility.

To confirm complete inactivation, cultured virus was diluted 1:10 into each candidate swab collection solution or water and subjected to heat-inactivation as described above. Inactivated virus was added to cultured Vero E6 cells under BSL3 conditions, and viral infection was assessed by scoring for cytopathic effect (CPE) after 3 days and 7 days. Cells inoculated with non-inactivated SARS-CoV-2 served as a positive control.

### RNA Purification using the Qiagen RNeasy kit

RNA was purified from 100 µl of each swab sample using the RNeasy Plus Mini Kit (Qiagen, Cat. #74136). 600 µl of buffer RLT was used in the first step, and RNA was eluted with 50 µl of water in the final step. The gDNA eliminator column step was omitted.

### RNA Purification using the QIAmp Viral RNA Mini kit

RNA was purified using the QIAmp Viral RNA Mini kit (Qiagen Cat. #52906) following the manufacturer’s instructions. Briefly, 140 µl of each sample was mixed with 560 µl of buffer AVL containing carrier RNA and incubated for 10 min at room temperature. After addition of 560 µl of 100% ethanol, the samples were passed through purification columns by centrifugation. The columns were washed sequentially with 500 µl of buffer AW1 and 500 µl of buffer AW2, and RNA was eluted using 40 µl of RNAse-free water.

### RNA Purification using the MagMax kit

RNA was purified from cultured virus diluted in DNA/RNA Shield using the MagMAX™ Viral RNA Isolation Kit (Thermo Fisher), following the protocol used for clinical samples in a CLIA-approved lab [26]. 450 µl of each sample was mixed with 10 µl of proteinase K, 10 µl of MS2 phage control, 265 µl of binding buffer, and 10 µl of magnetic beads, and the mixture was incubated at 65°C for 10 min. Beads were pelleted on a magnetic rack and washed once with 750 µl of wash buffer and twice with 500 µl of 80% ethanol, thoroughly resuspending and re-pelleting the beads at each wash step. Ethanol was thoroughly aspirated after the final wash step, and the beads were allowed to air dry at room temperature for 2 minutes. After addition of 25 µl of elution solution to each sample, the beads were thoroughly resuspended by agitation at 1400 rpm in a thermomixer (~3 min) and incubated for a total of 10 min at 65°C. Beads were pelleted again on a magnetic rack, and 20 µl of RNA eluate was withdrawn by pipetting.

### Isopropanol precipitation

Swab samples were inactivated using heat or DNA/RNA Shield as described above. 100 µl of each swab sample was mixed in 1.7 ml microcentrifuge tubes with 0.1 volumes of 3 M sodium acetate, pH 5.2, and 1 µl of 5 mg/ml linear acrylamide. Samples were then mixed with 1.1 volumes of isopropanol, incubated at −20°C for 30 min, and centrifuged at 16,000 *g* for 15 min at 4°C. Supernatants were aspirated, taking care not to disturb the pellets containing RNA. 1 ml of 75% ethanol was added to each sample, and samples were centrifuged again at 16,000 *g* for 5 min at 4°C. Supernatants were carefully but thoroughly aspirated. RNA was redissolved by adding 50 µl of water directly to each pellet and incubating for 10 min at 30°C.

### RT-qPCR with TaqPath master mix

RT-qPCR reactions with TaqPath master mix (Thermo Fisher) were assembled following the manufacturer’s instructions. For a 20 µl reaction, 5 µl of 4x TaqPath master mix was combined with 1.5 µl of SARS-CoV-2 (2019-nCoV) CDC N1, N2, or RNase P qPCR Probe mixture (Integrated DNA Technologies, Cat. #10006606), RNA sample, and water to a final volume of 20 µl. Volumes were divided by 2 for 10 µl reactions. RT-qPCR was performed on a BioRad CFX96 or CFX384 instrument with the following cycle: 1) 25°C for 2 min, 2) 50°C for 15 min, 3) 95°C for 2 min, 4) 95°C for 3 s, 5) 55°C for 30 s (read fluorescence), 6) go to step 4 for 44 additional cycles.

### Enzyme purification and BEARmix reactions

A detailed protocol for purification of Taq DNA polymerase and M-MLV reverse transcriptase and preparation of BEARmix can be found on GitLab: https://gitlab.com/tjian-darzacq-lab/bearmix.

In brief, a 4x buffer + dNTP mixture (“4xBEARbuffer+dNTPs”) was prepared containing the following components:

200 mM Tris-HCl, pH 8.4
300 mM KCl
12 mM MgCl_2_
40% trehalose
40 mM DTT
0.4 mM EDTA
4 mM each of dATP, dCTP, dGTP, dTTP

Separately, a 100x enzyme mixture (“100x BEAR enzymes”) was prepared containing 1.6 mg/ml of homemade Taq DNA polymerase and 0.17 mg/ml of homemade M-MLV reverse transcriptase in storage buffer (50 mM Tris-HCl, pH 8, 100 mM NaCl, 0.1 mM EDTA, 5 mM DTT, 0.1% Triton X-100, 50% glycerol).

For a 10 µl reaction, the following components were mixed on ice:

|  |  |
| --- | --- |
| 4x BEARbuffer+dNTPs | 2.5 μl |
| 100x BEAR enzymes | 0.1 μl |
| Primer/probe mixture (containing 6.7 μM of forward and reverse primers and 1.7 μM of TaqMan probe) | 0.75 μl |
| Sample (up to 6.65 μl) | X μl |
| Water | (6.65 - X) μl |
| Total volume | 10 μl |

The block of a qPCR machine was allowed to pre-heat to 50°C, and reactions were performed using the following cycle:

1. 50°C for 10 min
2. 95°C for 5 min
3. 95°C for 3 s
4. 55°C for 30 s; plate read
5. Goto 3, 44 additional times

The 95°C incubation in step 2 was extended to 10 min for the experiments shown in Fig 4 and to 10, 15, or 20 min for the hot-start Taq reactions in S7 Fig. A total of 50 rather than 45 cycles were used for the experiments in Fig 4A and S5 and S7 Figs.

### Quantification cycle (Cq) determination by the second-derivative method

Custom MATLAB code (available at https://gitlab.com/tjian-darzacq-lab/second-derivative-cq-analysis) was used to take the numerical second derivative of fluorescence intensity as a function of cycle number, averaged over a 3-cycle sliding window. If the second derivative peak was at the last cycle, then this was taken to be the Cq value. Otherwise, the Cq value was taken to be the center of the second derivative peak, as determined by fitting to a parabola. A user-selected second derivative cutoff was applied to all the samples within each experiment to distinguish amplification from non-amplification.

### Chemidoc Imaging of Plates

96-well plates and 8-well strips were imaged after PCR using a BioRad Chemidoc MP. The “fluorescein” preset was used to image FAM probe fluorescence with an integration time of 50 ms for Fig 7A-B and 100 ms for Fig 7C. For TaqPath reactions, the “rhodamine” preset with was used to image ROX loading control dye fluorescence with an integration time of 500 ms. The fluorescein and rhodamine channel images were overlaid in Fiji (https://imagej.net/Fiji) for Fig 7A-B. The tubes in Fig 7C are displayed using the built-in “Green Fire Blue” colormap in Fiji.

### Proteinase K activity assays

Proteinase K from a frozen aliquot stored at −20°C was diluted to 200 jg/ml in either water or Solution 2 and stored at room temperature or 4°C for 1 to 19 days. As a control, fresh PK was diluted to 200 jg/ml and kept on ice for 15 minutes prior to setting up the reactions. Each reaction contained 4 µl of PK solution, 2 µl of 1 mg/ml BSA in water, and 14 µl of either water or Solution 2. BSA was added last, and reactions were incubated for 1 hour at room temperature. Reactions were stopped after 1 hour by adding 4, µl of SDS loading buffer (200 mM Tris pH 6.8, 400 mM DTT, 10% BME, 8% SDS, 0.4% bromophenol blue, 40% glycerol) and heating the tubes at 95°C for 2 minutes. 12 µl of each reaction was separated on a 4-20% SDS-PAGE gradient gel (BioRad). The gel was fixed and stained with Flamingo Fluorescent protein stain (BioRad) following the manufacturer’s instructions and imaged on a Chemidoc (BioRad) imaging system. Undigested BSA migrates at ~66 kDa, while digestion products migrate below ~30 kDa.

### Preparation and analysis of contrived swab samples

Contrived swab samples were prepared under BSL3 conditions by mixing 3.2 µl of a 3.16 x 10^6^ PFU/ml viral stock (10^4^ PFU) or of a 1:10 dilution of this stock in 1x PBS (10^3^ PFU) to 50 µl of pooled human nasal fluid (Innovative Research, product # IRHUNF1ML). This mixture was then diluted into 1 ml of either 1x DNA/RNA Shield (Zymo Research), VCM with 0.4 mg/ml proteinase K (PK), or water with 0.4 mg/ml PK. Control samples were prepared with the same quantities of virus but without nasal fluid. Samples containing PK were incubated at 37°C for 30 min and then heat-inactivated at 75°C for 30 min. RNA was purified using the QIAmp Viral RNA Mini kit, as described above. Alternatively, 1 µl, 5 µl, or 13 µl of each sample was added directly to a BEARmix RT-qPCR. Reactions were performed as described above.

## Data Availability

Raw data from this manuscript are available upon request.

## Acknowledgments

We would like to thank Kaiser Permanente Healthcare and Fyodor Urnov for access to swab samples, Dirk Hockemeyer for helpful discussions, the Tjian-Darzacq lab for daily Zoom meetings and brainstorming, David Long (Medical University of South Carolina) for advice on master mix and hot-start Taq preparation, and Chips Hoai, Laura Flores, Anna Maurer, Doug Fox, and the UC Berkeley Environmental Health and Safety Office for assistance with biological safety and regulatory compliance.

## Financial Disclosure

The authors gratefully acknowledge funding from the Howard Hughes Medical Institute (Grant CC34430 to R.T.) This work was largely funded by Xavier Darzacq and Robert Tjian’s discretionary funds. T.G. is supported by a postdoctoral fellowship from the Jane Coffin Childs Memorial Fund for Medical Research.

## Author contributions

C.D. and T.G. designed and performed most of the experiments, under the supervision of R.T. and X.D., and T.G. prepared most of the figures. G.D. and T.G. developed and optimized the formulation of BEARmix, for which T.G. purified the enzymes. X.N. and E.V. performed all BSL3 procedures, under the supervision of S.S. M.E. performed BSA proteolysis experiments to evaluate the shelf life of proteinase K. A.A. assisted with initial purification of RNA from NP swab samples. T.G., M.E., and C.D. wrote the manuscript, and the remaining authors edited the manuscript.

## Notes

### Competing Interest Statement

The authors have declared no competing interest.

### Author Declarations

The UC Berkeley Office for Protection of Human Subjects concluded that our project does not meet the threshold definition of human subjects research set forth in Federal Regulations at 45 CFR 46.102(f).

